# The interpretation of clinical relevance in randomised clinical trials in patients with chronic low back pain: protocol for a meta-research study

**DOI:** 10.1101/2022.12.14.22283454

**Authors:** Tiziano Innocenti, Tim Schleimer, Stefano Salvioli, Silvia Giagio, Raymond Ostelo, Alessandro Chiarotto

**Affiliations:** Department of Health Sciences, Faculty of Science, Vrije Universiteit Amsterdam, Amsterdam Movement Sciences, The Netherlands; Department of Applied Health Sciences, Division of Physiotherapy, Hochschule für Gesundheit, Bochum, North Rhine-Westphalia, Germany; Department of Neuroscience, Rehabilitation, Ophthalmology, Genetics, Maternal and Child Health, University of Genoa, Genoa, Italy; Division of Occupational Medicine, IRCCS Azienda Ospedaliero-Universitaria di Bologna, Bologna, Italy; Department of Biomedical and Neuromotor Sciences (DIBINEM), Alma Mater Studiorum, University of Bologna, Bologna, Italy; Department of Epidemiology and Data Science, Amsterdam UMC, Location Vrije Universiteit, Amsterdam Movement Sciences research institute, The Netherlands; Department of General Practice, Erasmus MC, University Medical Center, Rotterdam, the Netherlands

## Abstract

**Background:** Chronic low back pain (cLBP) is one of the leading worldwide causes of disability, which accounts for large costs for healthcare systems and work productivity. Many treatment options are available for patients with cLBP and to determine the effectiveness of an intervention, several stakeholders (e.g. researchers, policymakers) should make a judgment about the statistical significance and clinical relevance of the results. Many parameters have been proposed to find a threshold of clinical relevance and the Minimal Important Difference (MID) has been used for a long time. However, the MID has many potential flaws: it is not intervention-specific, and it should be used to interpret a within-group change on an outcome measure rather than as a parameter to interpret a between-group effect. To overcome these issues, the smallest worthwhile effect (SWE) parameter has been proposed. The SWE captures the judgments of recipients of care; allows patients to weigh the benefits of treatment against the risks, costs, and side effects; and potentially provides estimates based on an intervention-control comparison. Specific values of the SWE have been calculated in patients with cLBP for pain intensity, physical functioning and time to recovery for physiotherapy (combining exercise and manual therapy) compared with no intervention. These values are expressed as a percentage of the smallest amount of patient-valued benefit that an intervention would require, compared to an alternative, to justify associated costs, risks, and other inconveniences.

**Objectives:** 1) To evaluate how authors of original, eligible, and published RCTs have interpreted the clinical relevance of the effect of physiotherapy compared to no-intervention on pain, physical functioning and time to recovery; 2) To reinterpret the clinical relevance of these between-group differences of these original, eligible and published RCTs based on the available SWE estimates for physiotherapy in cLBP compared to no intervention; 3) To evaluate, for descriptive purposes, whether the studies are adequately powered or underpowered considering the published SWE values and a power threshold of 80%.

**Methods:** A systematic search in Medline, PEDro, Embase and Cochrane CENTRAL from inception to November 30^th^ 2022 will be conducted. We will search for randomized controlled trials investigating the effectiveness of physiotherapy (combining exercise and manual therapy) as compared to no interventions in people with cLBP. Two authors will perform the data selection and extraction independently; a third reviewer will be involved in case of disagreement. We will compare the authors’ interpretation of results for statistical significance and clinical relevance with their results to determine if they meet their a-priori definitions. Then, we will perform a re-interpretation of the between-group differences for every individual trial based on SWE values published for cLBP. Lastly, we will calculate the power for each included study considering the alpha error used in the study, sample size and a between-group difference of 20% on the primary outcome.

**Ethics and dissemination:** A manuscript will be prepared and submitted for publication in an appropriate peer-reviewed journal upon study completion. We believe that the results of this investigation will be relevant to researchers paying more attention to the interpretation of the research results to translate clinical implications to key stakeholders (healthcare providers and patients).

**VERSION 2.0 – CHANGES:**

- **We changed the title replacing “meta-epidemiological” with “meta-research”**. This decision is made to reflect the absence of an inferential approach (e.g. regression analyses) in our study that usually characterizes meta-epidemiological studies, as highlighted in a recent publication (see: Puljak L. Caution is needed when describing a study design as meta-epidemiological. J Clin Epidemiol. 2022 Dec;152:326-327. doi: 10.1016/j.jclinepi.2022.10.017. Epub 2022 Oct 26. PMID: 36309145).
- **We replaced meta-epidemiological with “meta-research” in the whole text**.

## INTRODUCTION

Chronic low back pain (cLBP) is defined as pain, muscle tension, or stiffness lasting longer than 12 weeks^1^ and it is one of the worldwide leading causes of disability, which accounts for large costs for healthcare systems and work productivity^2,3^. Around 65% with LBP still report pain after one year^4^, and it affects physical, emotional and social functioning^3^.

Many treatment options are available for patients with cLBP^5^. Randomised controlled trials (RCT) provide the best study design yielding accurate estimates of the effectiveness of an intervention^6^. To determine the effectiveness of an intervention, various stakeholders (e.g. researchers, policy makers) should make a judgment about the statistical significance and clinical relevance of the results^7^.

Clinical relevance indicates whether the intervention should be used in clinical practice to improve healthcare outcomes, and it involves the interpretability of between-group differences^8^. Between-group differences represent the effect between two or more treatments to determine if the effect of the treatment is clinically relevant^8^. Many parameters have been proposed to find a threshold of for interpreting the clinical relevance of between-group differences^9,10^. Several terms have been used to identify a threshold for clinical relevance over the years^10,11^. It was first described as “Minimal Clinical Important Difference – MCID” in 1989 by Jaeschke et al.^9^. In 2005 the “C” was removed because it focused on the clinic rather than the patients’ experience^12^. Since its first definition, many other studies^13-16^ have described similar entities with slightly different names, but it is not always clear what construct these measurements are intended to measure^11^. Now, the minimal important difference (MID) is defined as “the smallest difference in score in the outcome of interest that informed patients or informed proxies perceive as important, either beneficial or harmful, and which would lead the patient or clinician to consider a change in the management”^12^.

Good estimates of between-group clinical relevance interpretation should satisfy two conditions, as argued by Barrett et al.^10^. The first is that they should be intervention-specific and must involve weighing the benefits of the intervention against its costs, risks, and side effects, as compared to a control intervention. The second is that the judgments about whether the benefits of intervention should be based on the perspective of patients who receive the intervention. In addition, these estimates should be expressed in terms of a between-group difference effect rather than a within-group change on a specific outcome measure, especially if they want to be used to inform the design and interpretation of clinical trials^8,17^.

The MID does not fully satisfy these aforementioned requirements: costs, risks, and other harms are not generally taken into account within the existing MID framework^10^, it is not intervention-specific, and it should be used to interpret a within-group change on an outcome measure rather than a between-group difference. To overcome these issues, in 2005, Barrett et al.^10,18^ described a methodological approach, the ‘‘benefit harm trade-off method,’’ to estimate the smallest worthwhile effect (SWE) of health interventions. The SWE approach captures the judgments of recipients of care; allows patients to weigh the benefits of treatment against the risks, costs, and side effects; and potentially provides estimates based on an intervention-control comparison. It is expressed as a percentage of the smallest amount of patient-valued benefit that an intervention would require to justify associated costs, risks, and other inconveniences^10^.

Two studies^19,20^ calculated SWE values for pain intensity, physical functioning and time to recovery for physiotherapy interventions (exercise and manual therapy) compared with no intervention for patients with cLBP. These studies consistently found that patients need to experience at least a 20% additional improvement in pain and physical functioning, and to speed up their recovery by 10 days to consider that the effect of physiotherapy is worthwhile given its costs, potential side effects, and inconveniences, as compared to receiving no intervention. Exploring how the authors of published RCTs on physiotherapy for cLBP have interpreted their results and reinterpreting the clinical relevance in light of the SWE values for between-group differences could be relevant to future research (e.g., to drive the sample size calculation in RCTs) and clinical practice (e.g., to make clinicians and patients able to better judge the clinical relevance of physiotherapy interventions in cLBP).

Therefore, the objective of our meta-research study are the following:

### Primary objectives

- To evaluate how authors of eligible RCTs have interpreted the clinical relevance of the between-group difference of physiotherapy interventions compared to no-intervention on pain, physical functioning and time to recovery;
- To re-interpret the clinical relevance of the between-group differences of the published RCTs based on the available SWE estimates for physiotherapy compared to no intervention in patients with cLBP ^19,20^.

### Secondary objectives

- To evaluate, for descriptive purposes, whether the studies are adequately powered or underpowered considering the published SWE values and a power threshold of 80%.

## MATERIAL AND METHODS

We will follow and adapt the PRISMA 2020 checklist^21^ for the reporting of this manuscript (or a specific reporting checklist for meta-research studies, if available at the time of reporting^22^).

### Data source and search strategies

For retrieving eligible articles, a literature search on the following electronic databases will be conducted from inception to November 30th, 2022: Medline (through the PubMed interface), PEDro, Embase (Embase.com), and Cochrane CENTRAL (www.cochranelibrary.com). In addition to the electronic database search, other potentially relevant studies will be searched in the reference lists of the included articles and recent systematic reviews on the topic^1^, in all relevant grey literature sources such as conference proceedings, OpenGray, and Google Scholar. The search will be re-run before the final analyses to retrieve any additional records.

Medical subjects’ headings (MeSH) and all relevant free text words will be combined using the boolean operators (e.g. AND, OR): (1) condition (chronic low back pain); (2) intervention (exercise and manual therapy); (3) study design (Randomized Controlled Trial, Controlled Clinical Trial). The Cochrane MEDLINE sensitivity-maximizing RCT filter will be used to ensure the maximum sensitivity to our search strategy since it has very high sensitivity and a slightly better precision relative to more sensitive filters^23^.

No year restrictions will be applied. We will include studies written in English, French, Italian, German, Dutch, Spanish and Portuguese. If we find eligible articles in other languages (e.g. Chinese or Japanese), we will try to translate them using online web services or look for native-speaker readers to perform their assessment. Then, if we cannot reach a suitable translation or find a native speaker, we will exclude them from our sample.

A detailed search strategy will be created for each electronic database (see Appendix 1).

### Eligibility criteria

To be consistent with the criteria used in the validation studies for the SWE^19,20^, the following inclusion criteria were applied to the retrieved literature:

1. conducted in adult patients (>18 years old) with non-specific cLBP (defined as LBP lasting for more than 12 weeks)
2. The intervention consisted of a combination of the following physiotherapy treatment components: any exercise therapy modality (e.g. but not limited to resistance exercise, strengthening exercise, aerobic exercise) AND any form of manual therapy (e.g. but not limited to muscle energy techniques, massage therapy, spinal manipulative therapy). Other treatments (e.g. education, psychologically informed physiotherapy, electrotherapy or acupuncture) can be included if they represent only a minor part of the intervention (i.e. <=25% of the treatment time/sessions).
3. The control group is ‘no intervention’ (e.g., waiting list or total absence of interventions). Regardless of the name given to the comparison by the original authors, we will include it if no active treatment is given to the patients. If, for example, authors describe usual care as “participants were instructed to follow their normal schedule of medications and physical activity”, we will include it. If only instructions were given without “an active role of the health care provider, “ we will include it.
4. at least one of the outcomes is pain intensity, physical functioning or time to recovery
5. RCT design and cross-over design (only the data before the crossover will be included), (6) the follow-up time point is short-term, defined as ≤ 12 weeks (since the study by Ferreira^20^ and Christiansen^19^ focused on 2 and 6 weeks, respectively).

The following exclusion criteria will be used:

1. patients undergoing back surgeries in the last year
2. patients with severe psychiatric co-morbidities that impede participation
3. other treatments (e.g., pain medication, injections) combined with physiotherapy. RCTs including a “mixed” population of patients with non-specific LBP and other musculoskeletal disorders (e.g., specific LBP, neck pain, acute LBP) will be included only if at least 75% of the patients displayed chronic LBP.

### Data selection

First, the titles and abstracts of the retrieved studies will be divided into two sets. Each set will be screened independently by two pairs of reviewers (TI, TS and SS, SG) to evaluate whether they meet the eligibility criteria. Second, the full texts of the potentially eligible studies will be downloaded and assessed for eligibility by the same two pairs. If the full text cannot be retrieved, we will contact the authors with a maximum of three emails. A consensus meeting will be held to determine agreement on the selection; in case of disagreement, a third reviewer (AC) will decide on inclusion. The reasons for the full-text exclusions will be recorded. The study selection process will be summarised through the PRISMA flowchart^21^. EndNote^24^ will be used to remove duplicates and manage the bibliography. Rayyan QCRI systematic review software^25^ will be used to carry out the selection process.

### Data extraction

The data will be extracted by two independent reviewers (TI and TS) and double-checked by a third reviewer (SS) to ensure the precision and quality of the retrieved information. The following data will be extracted from the included studies through a standardised extraction Excel sheet: design, study setting (author, year of publication, country, and language), in- and exclusion criteria, sample size, patient characteristics (e.g. age, sex, pain, physical functioning and time to recovery at baseline), type of intervention (including intervention characteristics such as frequency, and duration), outcomes measures used (for physical functioning, pain and time to recovery), effect sizes, cut-off used for statistical significance and clinical relevance, the description of the framework used (e.g. MID, SWE), follow-up time point.

Regarding statistical significance, the authors’ a-priori definition (e.g. two side p-value < 0.05) will be searched in the text, and the alpha value will be used for the sample size calculation. For clinical relevance, it will be checked how the authors a-priori defined the MID or SWE for the between-group differences; these parameters will be searched in the article text and the sample size calculation (either in the study protocol when available or the full-text), looking at the difference they used to calculate it.

The following predefined decision rules will be used to select data from trials to prevent selective inclusion: (a) where trialists reported both unadjusted and adjusted values for the same outcome; adjusted values will be extracted; (b) where trialists reported data analysed; based on the intention-to-treat (ITT) sample and another sample (e.g. per-protocol, as-treated), ITT-analyzed data will be extracted.

Missing data or additional details will be gathered by contacting the corresponding author of included studies with a maximum of three emails. For Cross-over trials, only the data before the crossover will be extracted and included in the analyses.

#### Data analysis

Descriptive data of the included studies will be reported in tables as mean (standard deviations) or median (interquartile range) values. The following descriptive analyses will be performed:

### Primary analyses

- We will compare the authors’ interpretation of results for statistical significance and clinical relevance with their results to determine if they meet their a-priori definitions. If the authors do not specify any clinical relevance or statistical significance threshold, we will use the difference hypothesised for sample size calculation. If the authors do not report any threshold or the sample size calculation, we will report the lack of information.
- We will determine if the author’s a priori-definition of clinical relevance meets the SWE threshold.
- We will re-interpret the clinical relevance, comparing the between-group differences for every individual trial based and the published SWE values ^19,20^. For pain intensity and physical functioning, the results will be considered clinically relevant in case of a 20% more improvement from baseline when comparing physiotherapy to no intervention. For time to recovery, in case of 10 days or more. To retrieve the percentage of between-group difference, we will calculate it by comparing the between-group difference with the mean baseline value of the study for each outcome.

### Secondary analysis

- We will calculate the power for each included study considering the alpha error used in the study, sample size and a between-group difference of 20% (the SWE values already published) on the primary outcome. In this way, each study will be classified as the following:
  - Underpowered if the power achieved is less than 80%.
  - Well-powered if the difference is 80% or above.
  Pilot trials evaluating the feasibility of a future large RCT will be excluded from this analysis.

SPSS software (IBM SPSS Statistics for Macintosh, Version 28.0. Armonk, NY: IBM Corp) will be used for the analyses. G*Power software will be used for a posteriori power calculation^26^.

## Data Availability

All data produced in the present work will be contained in the final manuscript

## ETHICS AND DISSEMINATION

This study does not require an ethics review as we will not collect personal data; it will summarise information from publicly available studies.

A manuscript will be prepared and submitted for publication in an appropriate peer-reviewed journal upon study completion. The study findings will be disseminated at a relevant (inter)national conference. All results of this meta-research study will also be announced at (inter)national scientific events in musculoskeletal rehabilitation and research methods. The results of this investigation will be relevant to researchers paying more attention to the synthesis of the evidence to translate clinical implications to key stakeholders (healthcare providers and patients).

## AUTHOR CONTRIBUTIONS

TI, AC, and RO conceived and designed the study protocol. TI, SS, TS, SG, RO and AC were involved in conceptualising the study objectives and providing input into study selection criteria and plans for data extraction. All the authors, including TI, TS, SS, SG, RO and AC, approved the final version of the protocol.

### APPENDIX 1

#### Search strategies for the literature search

| Database | Strategy |
| --- | --- |
| Pubmed | **#1** "Back Pain"[MeSH Terms] OR back pain[All Fields] OR "lbp"[All Fields] OR "Back Injuries"[MeSH Terms] OR "back injury"[All Fields] OR "Lumbar Vertebrae"[MeSH Terms] OR "lumbar spine"[All Fields] OR "Sacroiliac Joint"[MeSH Terms] OR "sacroiliac pain"[All Fields] OR "Zygapophyseal Joint"[MeSH Terms] OR "facet pain"[All Fields] OR "Intervertebral Disc Displacement"[MeSH Terms] OR "disc pain"[All Fields] |
|  | **#2** "physical therapy modalities"[MeSH Terms] OR "physiotherapy"[All Fields] OR "Physical Therapists"[MeSH Terms] OR "physiotherapist"[All Fields] OR "exercise therapy"[MeSH Terms] OR "exercise therapy"[All Fields] OR "Physical Therapy Department, Hospital"[Mesh] OR "manipulation, osteopathic"[MeSH Terms] OR "manipulation, spinal"[MeSH Terms] OR "manipulation, chiropractic"[MeSH Terms] OR "orthopedic manipulation"[All Fields] OR "osteopathic manipulation"[All Fields] OR "spinal manipulation"[All Fields] OR "chiropractic manipulation"[All Fields] OR "musculoskeletal manipulations"[All Fields] OR "spinal manipulative therapy"[All Fields] OR "manual therapy"[All Fields] |
|  | **#3** ("randomized controlled trial"[Publication Type] OR "controlled clinical trial"[Publication Type] OR "randomized"[Title/Abstract] OR "placebo"[Title/Abstract] OR "drug therapy"[MeSH Subheading] OR ("randomly"[Title/Abstract] OR "trial"[Title/Abstract] OR "groups"[Title/Abstract])) NOT ("animals"[MeSH Terms] NOT "humans"[MeSH Terms]) |
|  | **#1 AND #2 AND #3** |
| PEDro | Title and abstract: "Low back pain"<br>Body part: lumbar spine, sacro-iliac joints or pelvis<br>Method: clinical trial |
| Embase | **#1** 'back pain'/exp OR 'back pain' OR 'lbp' OR 'back injuries'/exp OR 'back injuries' OR 'lumbar spine'/exp OR 'lumbar spine' OR 'sacroiliac pain'/exp OR 'sacroiliac pain' OR 'facet pain' OR 'intervertebral disc displacement'/exp OR 'intervertebral disc displacement' |
|  | **#2** 'physical therapy'/exp OR 'physical therapy' OR 'physiotherapy'/exp OR 'physiotherapy' OR 'exercise'/exp OR 'exercise' OR 'manipulation'/exp OR 'manipulation' OR 'manual therapy'/exp OR 'manual therapy' |
|  | **#3** 'low back pain'/dm AND 'randomized controlled trial'/de AND 'article'/it |
|  | **#1 AND #2 AND #3** |
| COCHRANE Central | **#1** back pain OR lbp OR lumbar spine OR sacroiliac pain OR facet pain |
|  | **#2** physical therapy modalities OR physiotherapy OR exercise therapy OR manipulation OR manual therapy |
|  | **#1 AND #2** |

